# Covid-19 Testing, Hospital Admission, and Intensive Care Among 2,026,227 United States Veterans Aged 54-75 Years

**DOI:** 10.1101/2020.04.09.20059964

**Authors:** Christopher T. Rentsch, Farah Kidwai-Khan, Janet P. Tate, Lesley S. Park, Joseph T. King, Melissa Skanderson, Ronald G. Hauser, Anna Schultze, Christopher I. Jarvis, Mark Holodniy, Vincent Lo Re, Kathleen M. Akgün, Kristina Crothers, Tamar H. Taddei, Matthew S. Freiberg, Amy C. Justice

## Abstract

**Importance:** Severe acute respiratory syndrome coronavirus 2 (SARS-CoV-2) infection causes coronavirus disease 2019 (Covid-19), an evolving pandemic. Limited data are available characterizing SARS-Cov-2 infection in the United States.

**Objective:** To determine associations between demographic and clinical factors and testing positive for coronavirus 2019 (Covid-19+), and among Covid-19+ subsequent hospitalization and intensive care.

**Design, Setting, and Participants:** Retrospective cohort study including all patients tested for Covid-19 between February 8 and March 30, 2020, inclusive. We extracted electronic health record data from the national Veterans Affairs Healthcare System, the largest integrated healthcare system in the United States, on 2,026,227 patients born between 1945 and 1965 and active in care.

**Exposures:** Demographic data, comorbidities, medication history, substance use, vital signs, and laboratory measures. Laboratory tests were analyzed first individually and then grouped into a validated summary measure of physiologic injury (VACS Index).

**Main Outcomes and Measures:** We evaluated which factors were associated with Covid-19+ among all who tested. Among Covid-19+ we identified factors associated with hospitalization or intensive care. We identified independent associations using multivariable and conditional multivariable logistic regression with multiple imputation of missing values.

**Results:** Among Veterans aged 54-75 years, 585/3,789 (15.4%) tested Covid-19+. In adjusted analysis (C-statistic=0.806) black race was associated with Covid-19+ (OR 4.68, 95% CI 3.79-5.78) and the association remained in analyses conditional on site (OR 2.56, 95% CI 1.89-3.46).

In adjusted models, laboratory abnormalities (especially fibrosis-4 score [FIB-4] >3.25 OR 8.73, 95% CI 4.11-18.56), and VACS Index (per 5-point increase OR 1.62, 95% CI 1.43-1.84) were strongly associated with hospitalization. Associations were similar for intensive care. Although significant in unadjusted analyses, associations with comorbid conditions and medications were substantially reduced and, in most cases, no longer significant after adjustment.

**Conclusions and Relevance:** Black race was strongly associated with Covid-19+, but not with hospitalization or intensive care. Among Covid-19+, risk of hospitalization and intensive care may be better characterized by laboratory measures and vital signs than by comorbid conditions or prior medication exposure.

**Key Points:** *Question:* What are the demographic and clinical characteristics associated with testing positive for coronavirus 2019 (Covid-19+), and among Covid-19+ subsequent hospitalization and intensive care among Veterans in the United States?

*Findings:* In this retrospective cohort study of 2,026,227 Veterans aged 54-75 years and active in care, 585/3,789 (15.4%) tested Covid-19+. Black race was strongly associated with Covid-19+, but not with hospitalization or intensive care. Among Covid-19+, laboratory abnormalities and a summary measure of physiologic injury were strongly associated with hospitalization and intensive care.

*Meaning:* Racial differences in testing positive for Covid-19 may be an underestimate of the general population as racial health disparities in the Veterans Affairs Healthcare System tend to be smaller than in the private sector. Risk of hospitalization and intensive care may be better characterized by laboratory measures and vital signs than by comorbid conditions or prior medication exposure.

## Introduction

Severe acute respiratory syndrome coronavirus 2 (SARS-CoV-2) infection causes coronavirus disease 2019 (Covid-19) and is an evolving pandemic. Limited data are available characterizing SARS-Cov-2 infection in the United States. Unadjusted analyses restricted to Covid-19 cases in China,^1-5^ Italy,^6^ and the United States^7,8^ suggest that older age, diabetes, chronic obstructive lung disease (COPD), hypertension, vascular disease, renal disease, and liver disease are associated with more severe disease. Further, while some have speculated that use of angiotensin converting enzyme inhibitor (ACE), angiotensin II receptor blockers (ARB), and nonsteroidal anti-inflammatory drugs (NSAID) may exacerbate disease,^9,10^ no analysis of this question has been published.

The Department of Veterans Affairs (VA) is the largest integrated healthcare system in the United States. All care is recorded in a national electronic health record with daily uploads into a central data repository. As a result, it is possible to extract data on patients tested for Covid-19, including outpatient and inpatient records, laboratory values, and pharmacy fill/refill data. When a well-characterized longitudinal cohort is supplemented with Covid-19 testing data, it is possible to answer important questions rapidly using validated methods.

The VA Birth Cohort includes all Veterans born between 1945 and 1965, over 2 million living individuals aged 54-75 years,^11,12^ a demographic at particularly high risk of adverse outcomes from Covid-19.^1-4^ Using unadjusted and adjusted analyses, we consider a wide range of factors either associated with testing positive for Covid-19 and subsequent hospitalization and intensive care in the national VA system as of March 30, 2020.

## Methods

### Data Source

Using data from the VA national Corporate Data Warehouse on members of the VA Birth Cohort, we identified patients tested for Covid-19 from date of first recorded VA test on February 8, 2020 through March 30, 2020. Available data included demographics, outpatient and inpatient encounters, diagnoses, laboratory results, vital signs, health factors (e.g., smoking and alcohol health behaviors), and pharmacy dispensing records.

VA Birth Cohort was approved by the Institutional Review Boards of VA Connecticut Healthcare System and Yale University. It has been granted a waiver of informed consent and is Health Insurance Portability and Accountability Act compliant.

### Data Collection

We selected previously validated cohort characteristics and those that have been evaluated in prior Covid-19 reports.^1,13^ Baseline was defined as the date of specimen collection for the Covid-19 test unless testing occurred during hospitalization, in which case it was date of admission. Demographics included age at baseline, sex, race/ethnicity, and rural/urban residence. Residence was defined using geographic information system coding based upon established criteria.^14^

### Main Study Outcomes

We examined three outcomes: 1) testing positive for SARS-CoV-2 (Covid-19+), 2) hospitalization, and 3) admission to an intensive care unit (ICU). We used VA inpatient bed section codes 12 (medical) and 13 (cardiac) to identify ICU admission.

### Covid-19 tests

We identified Covid-19 tests conducted in the VA using text searching of laboratory results containing terms consistent with SARS-CoV-2 or Covid-19. If a patient had more than one test and all were negative we selected first negative, otherwise we used date of first positive. Patients for whom results were pending (n=93) or inconclusive (n=33) were excluded. Nearly all tests utilized nasopharyngeal swabs, 1% were from other sources. Testing was performed in VA, state public health and commercial reference laboratories using emergency use authorization approved SARS-CoV-2 assays.

### Comorbidity

We extracted diagnostic codes for asthma, cancer, COPD, chronic kidney disease, diabetes mellitus, hypertension, liver disease, vascular disease, and alcohol use disorder (definitions provided in **eTable 1**). We used a validated algorithm to capture smoking status derived from health factors.^15^

### Pharmacy Data

We collected pharmacy fills for ACE/ARBs, chemotherapy and immunosuppressive drugs, and prescription NSAIDs and determined which medications were active in the year prior to testing. Exposure windows for NSAIDS ended 14 days prior to baseline to minimize the potential of protopathic bias. Exposure windows for other medications not used to treat Covid-19 symptoms ended three days prior to baseline.

### Vital Signs, Clinical Laboratory Data, and a Summary Measure of Physiologic Injury

Vital signs measured within two days of baseline included body mass index (BMI), oxygen saturation, pulse, systolic blood pressure, and temperature. We chose laboratory findings closest to baseline within a year prior or up to one week after baseline. Measures included alanine aminotransferase, albumin, aspartate aminotransferase, creatinine, estimated glomerular filtration rate,^16^ fibrosis-4 score (FIB-4),^17^ hemoglobin, platelet count, total white blood cell count, and lymphocyte count. We calculated a validated composite measure of physiologic injury (VACS Index) which includes age, BMI, and all previously mentioned laboratory measures save lymphocyte count^18,19^ (details in **eMethods**).

### Statistical Analysis

We evaluated characteristics of patients undergoing Covid-19 testing, and among Covid-19+, factors associated with hospital admission and intensive care, using chi-square, Fisher’s exact, and Wilcoxon rank-sum tests, as appropriate. For bivariate comparisons, statistical significance reflects complete case analysis. When modeling Covid-19+, we restricted analyses to factors available when initially evaluating a patient (i.e., demographic data, comorbid conditions, medication history, health behaviors, and vital signs). Because age, black race, ACE/ARB use, and NSAID use are of special interest, we included them in all multivariable models. Otherwise, variables significant at p<0.05 in unadjusted analyses were included in the multivariable models. When modeling hospital admission and intensive care, we compared C-statistics for models including individual laboratory values to a model including VACS Index. In post hoc analyses we explored the association between black race and Covid-19+ with a multivariable model conditioned on site, among sites having at least five positive tests.

We report missing data for each variable. We used multiple imputation to impute missing laboratory measures, vital signs, and smoking status. The imputation model included outcomes and all covariates. Estimates from regressions performed on 10 imputed data sets were combined using Rubin’s rules.^20^ Analyses were performed using SAS version 9.4 (SAS Institute Inc., Cary, NC, USA) and Stata version 14.2 (StataCorp, LLC., College Station, TX). We used R version 3.6.3 to map Covid-19 cases in the VA system overall and those captured in the VA Birth Cohort.

## Results

In the year prior to the Covid-19 outbreak, the VA Birth Cohort included 2,026,277 living individuals: 1,866,256 (92.1%) men and 159,971 (7.9%) women. The cohort includes 1,369,454 (67.6%) white, 402,295 (19.9%) black, 106,639 (5.3%) Latinx, and 147,839 (7.3%) other or unknown race/ethnicity. More than a third of the subjects (745,284 or 36.8%) were 70-75 years of age, 23.3% (n=472,732) were 65-69 years old, 20.1% (n=407,900) were 60-64 years old, and 19.8% (n=400,311) were 54-59 years old. Of these, 3,789 individuals have been tested for Covid-19 (18.7 per 10,000 persons) since February 8, 2020 through March 30, 2020.

Testing per 10,000 persons varied by race, sex, age, and residence (p<0.001 for all). Black Veterans were more likely to be tested than white Veterans (28.0 versus 15.6). Women were more likely to be tested than men (23.3 versus 18.3). Testing generally decreased with age (age 54-59: 21.5; age 60-64: 22.2; age 65-69: 18.6; and age 70-75: 15.4). Veterans living in urban settings were more likely to be tested than those in rural settings (39.2 vs. 5.8).

Among those tested, median age was 65.7 years (**Table 1**), 90.2% were male, 29.7% were black, and 81.1% lived in urban settings. Common comorbid conditions were hypertension (65.0%), diabetes mellitus (37.8%), vascular disease (28.9%), COPD (26.2%), and alcohol use disorder (13.9%). Receipt of ACE/ARBs (40.5%) or NSAIDs (30.5%) was common. Among those tested, 42.3% were current smokers, 40.8% were obese (BMI >30 kg/m^2^), 7.7% were febrile (≥100.4°F), 13.1% were hypoxic (oxygen saturation ≤ 93%), and 35.4% were tachycardic (pulse ≥90 beats per minute).

**Table 1.**
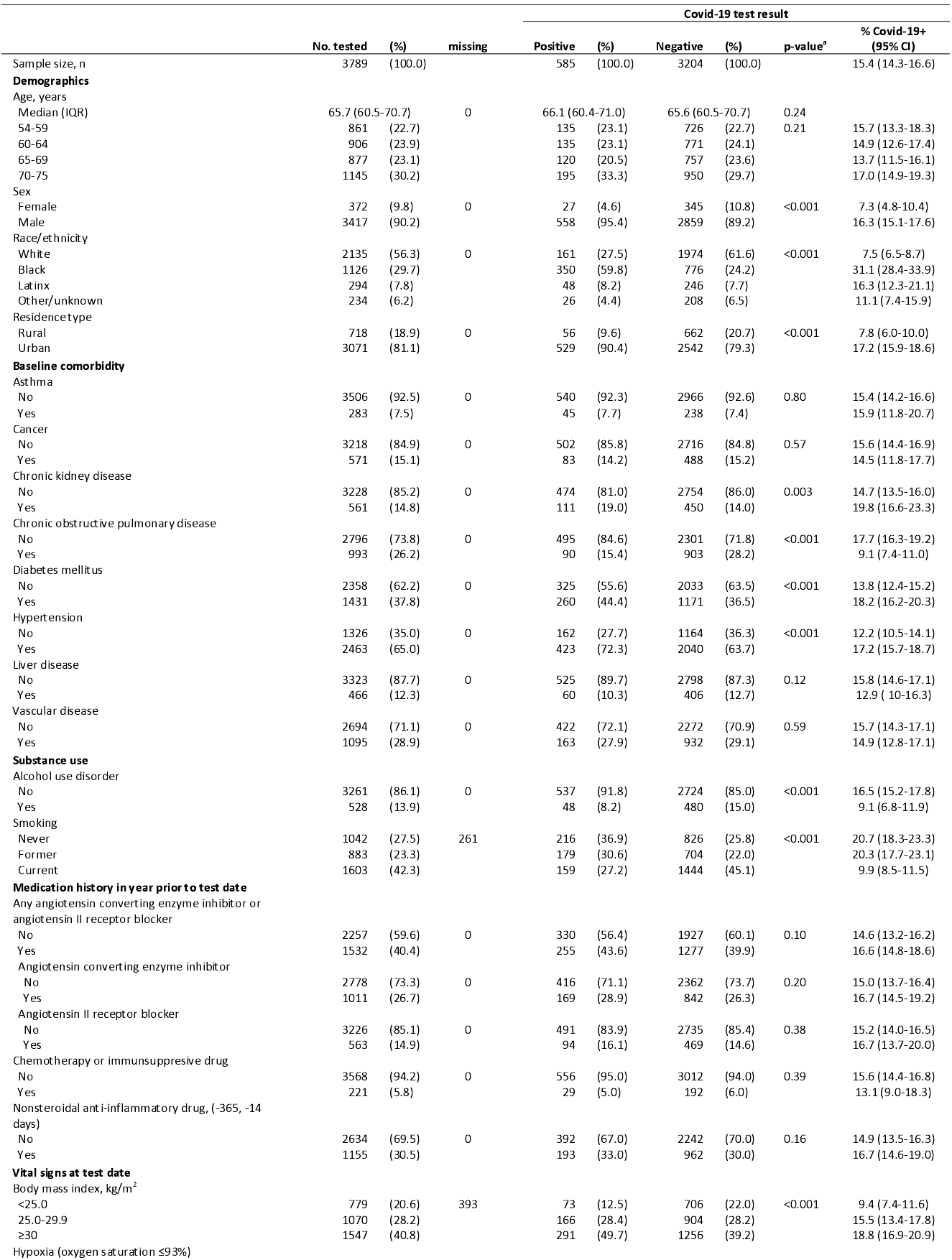
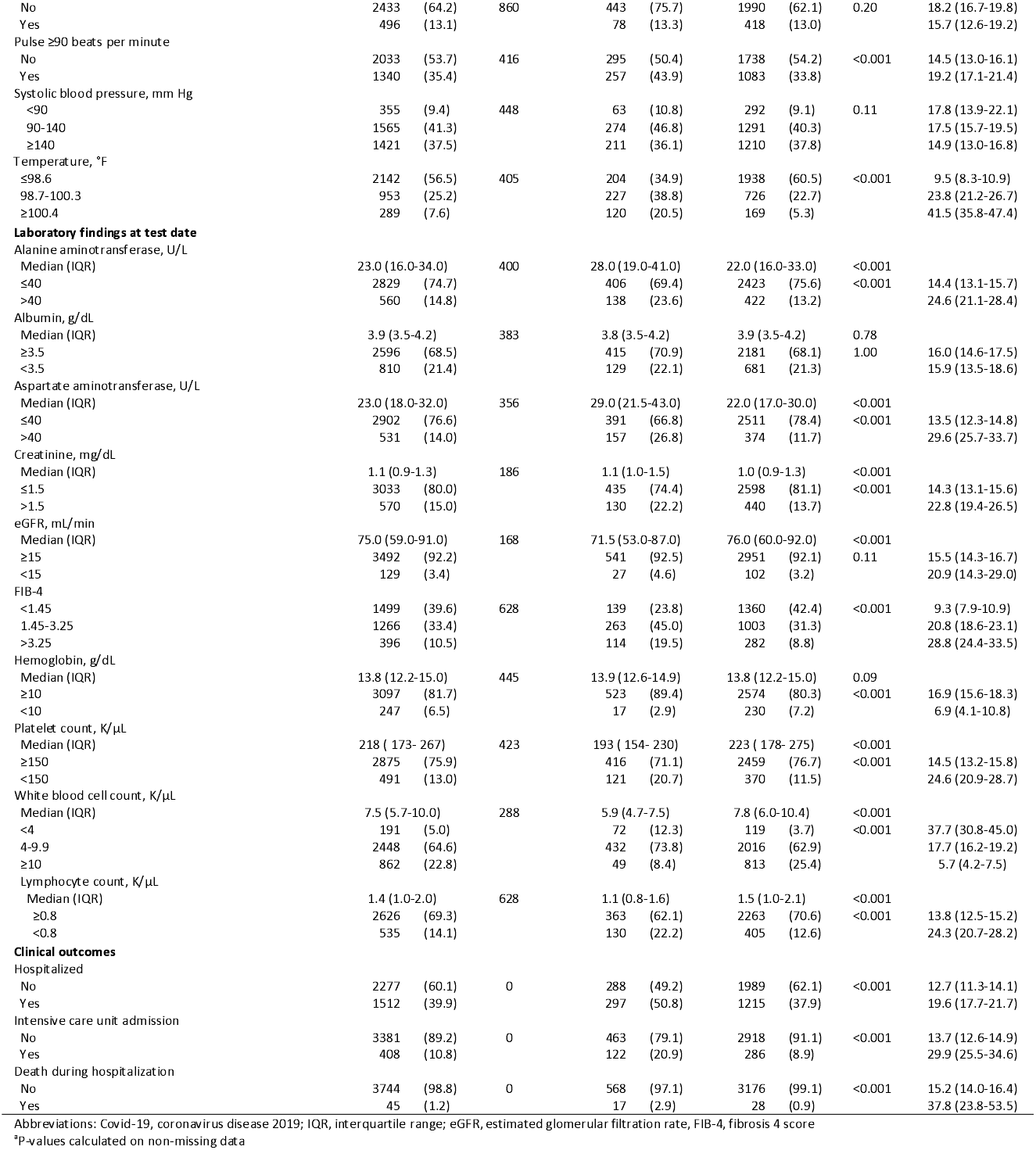
Characteristics of patients tested for Covid-19 among all patients aged 54-75 years in the Veterans Health Administration as of March 30, 2020

### Testing Positive vs. Negative for Covid-19

Of the 3,789 patients tested in the VA Birth Cohort, 585 (15.4%, 95% CI 14.3-16.6) were Covid-19+, representing approximately half (585/1244, 47%) of all Covid-19+ patients in the VA as of March 30, 2020 (**Figure 1a** and **eFigure 1**). In unadjusted analyses, factors associated with Covid-19+ (**Table 1**) included male sex, black race, urban residence, chronic kidney disease, diabetes, and hypertension (all p<0.003). Smoking, COPD, and alcohol use disorder were associated with a lower probability of a positive test (all p<0.001). No medication exposure was associated with a positive test. Vital signs associated with Covid-19+ included higher BMI, tachycardia, and higher temperature (all p<0.001). All laboratory values were associated with Covid-19+ (all p<0.001). Composite variables, eGFR and FIB-4, were also strongly associated (both p<0.001).

**Figure 1.**
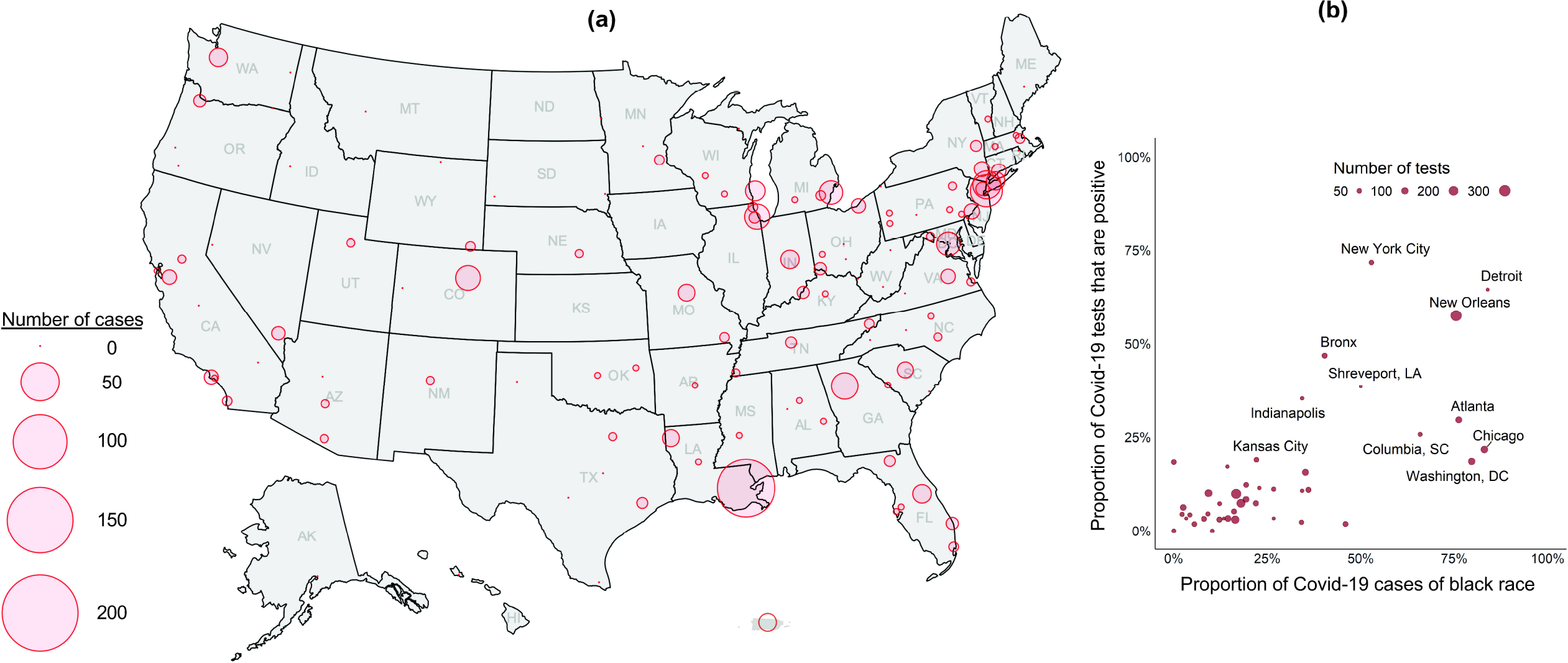
Distribution of Covid-19 cases in the Veterans Birth Cohort as of March 30, 2020 (a) Shown is the distribution of 585/1244 (47%) Covid-19 cases in the Veterans Health Administration captured in the Veterans Birth Cohort as of March 30, 2020 and included in the current study. (b) Shown is the proportion of Covid-19 test results that are positive by the proportion of Covid-19 cases of black race by site of care.

In multivariable analyses (**Table 2**, C-statistic=0.806), black race (OR 4.68,8 95% CI 3.79-5.78), male sex (OR 3.17, 95% CI 2.03-4.94), urban residence (OR 1.60, 95% CI 1.17-2.20), higher temperature (OR 1.70, 95% CI 1.58-1.84 per 1°F), lower systolic blood pressure (OR 1.44, 95% CI 1.16-1.78), and prior use of NSAIDS (OR 1.27, 95% CI 1.02-1.58) were associated with increased likelihood of Covid-19+. Current smoking (OR 0.45, 95% CI 0.35-0.57), alcohol use disorder (OR 0.58, 95% CI 0.41-0.83), and COPD (OR 0.67, 95%CI 0.50-0.88) were associated with decreased likelihood of Covid-19+. Results were similar in complete case analysis (**eTable 2**).

**Table 2.**
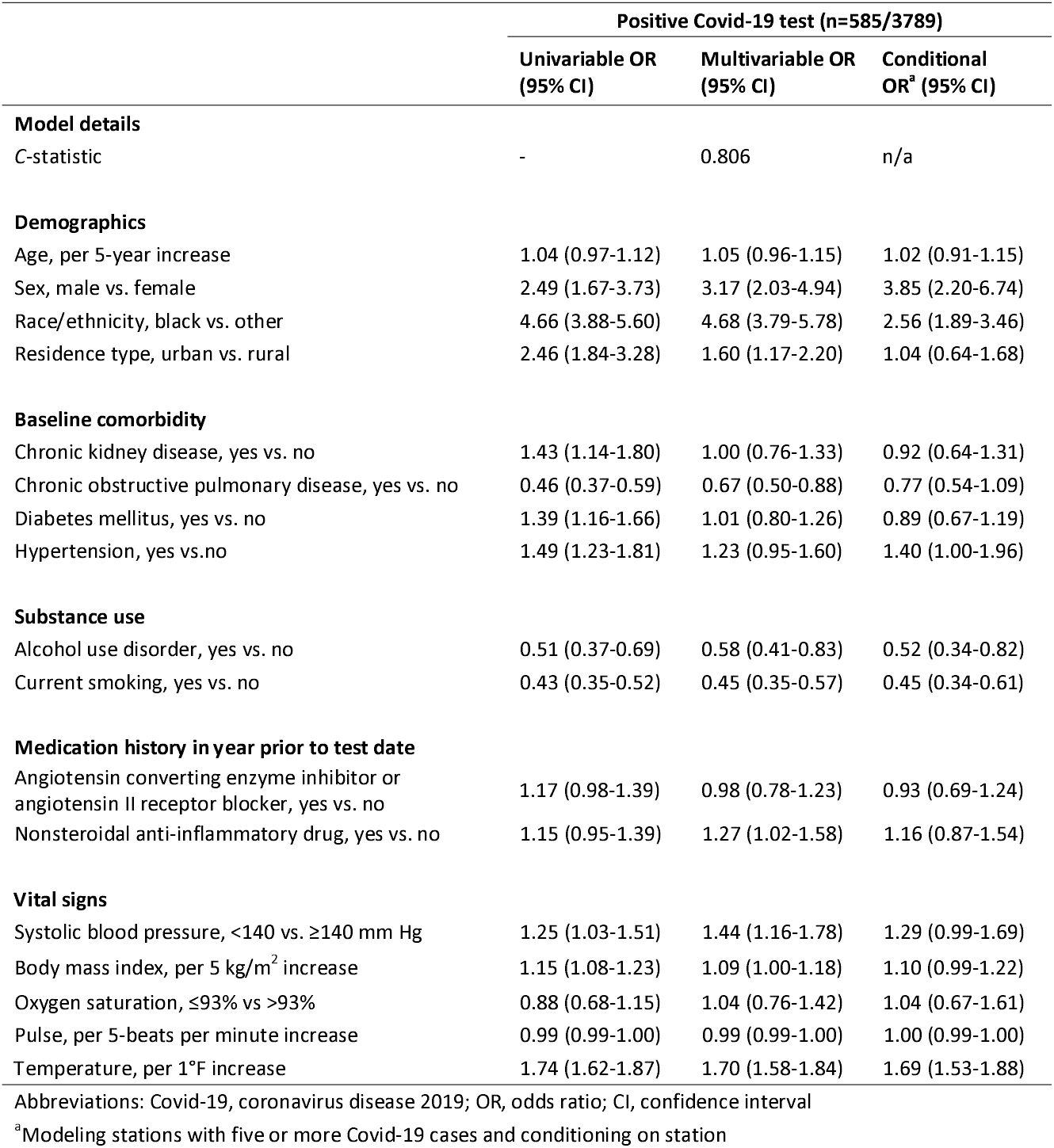
Crude and adjusted associations with testing positive for Covid-19 as of March 30, 2020

In *post hoc* analyses, we observed that black Veterans were more likely to be tested at sites with higher Covid-19 prevalence (**Figure 1b**). A model conditional on site (**Table 2**) reduced the association with black race (OR 2.56, 95% CI 1.89-3.46) and increased the association with male sex (OR 3.85, 95% CI 2.20-6.74). Associations with other factors were consistent with unconditional estimates.

### Risk Factors for Hospitalization and Intensive Care

Among 585 Covid-19+ patients, 297 (50.8%, 95% CI 46.6-54.9%) were hospitalized and 122 (20.9%, 95% CI 17.6-24.4%) received intensive care. In bivariate analyses, age, chronic kidney disease, COPD, diabetes, hypertension, vascular disease, ACE/ARB exposure, and decreased oxygen saturation, and elevated temperature were associated with hospitalization and intensive care (all p<0.05, **Table 3a and Table 3b**). All laboratory abnormalities investigated were associated with hospitalization and intensive care (all p<0.05). Median VACS Index scores varied substantially between those hospitalized versus not hospitalized (78.7 vs. 66.2, p<0.001) and between those receiving and not receiving intensive care (82.0 vs. 69.4, p<0.001).

**Table 3a.**
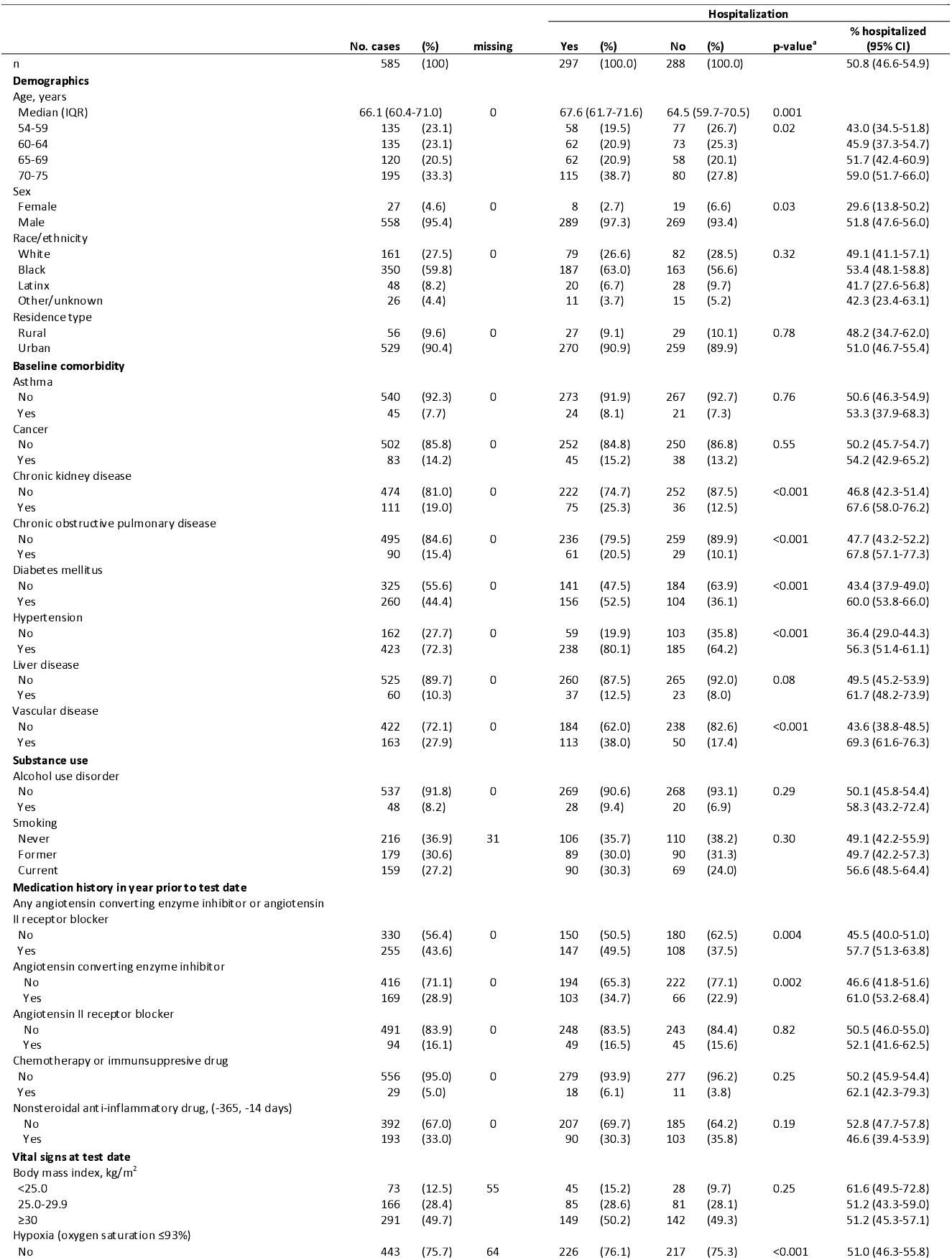
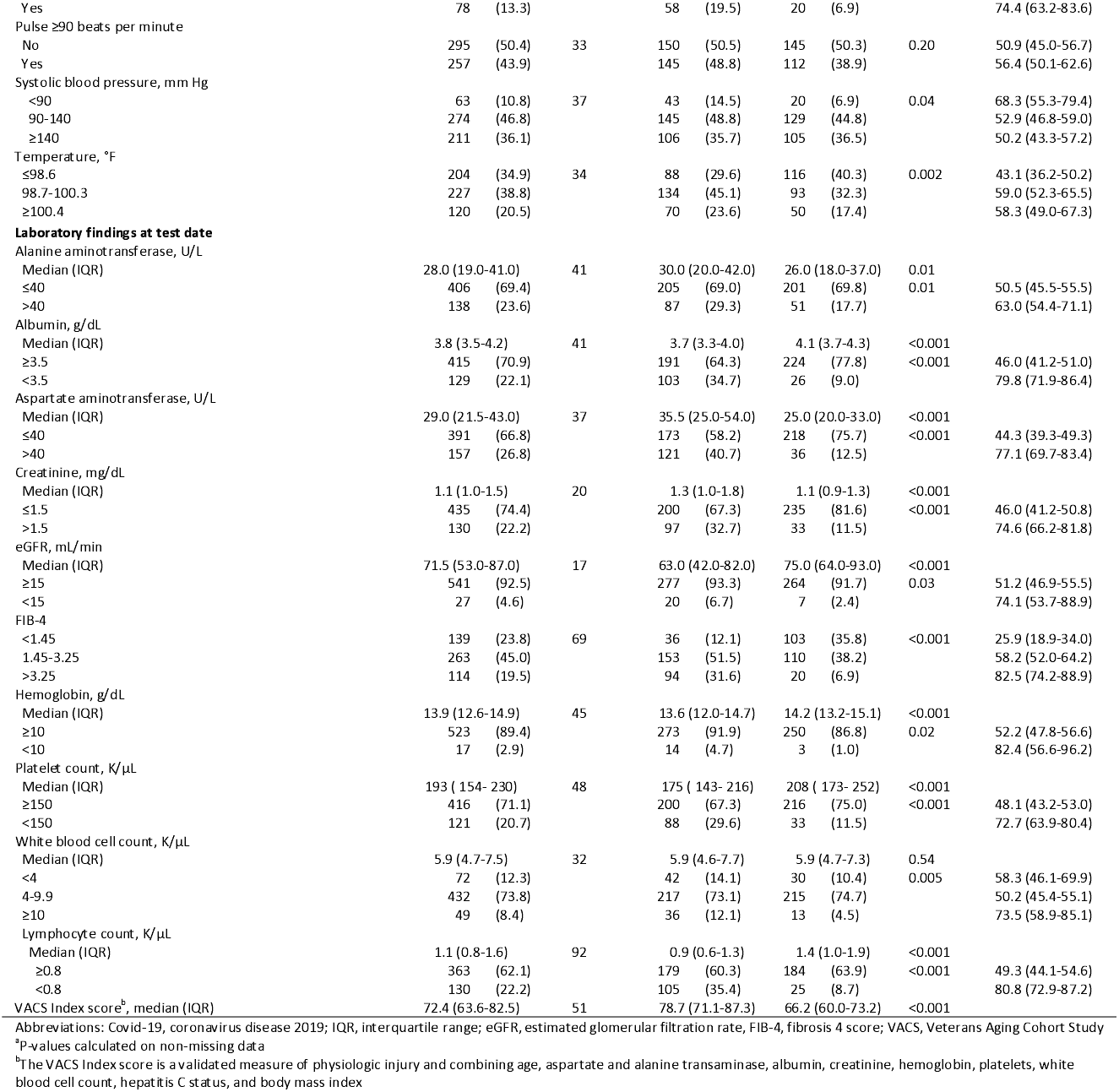
Associations with hospitalization among Covid-19 cases aged 54-75 years in the Veterans Health Administration as of March 30, 2020

**Table 3b.**
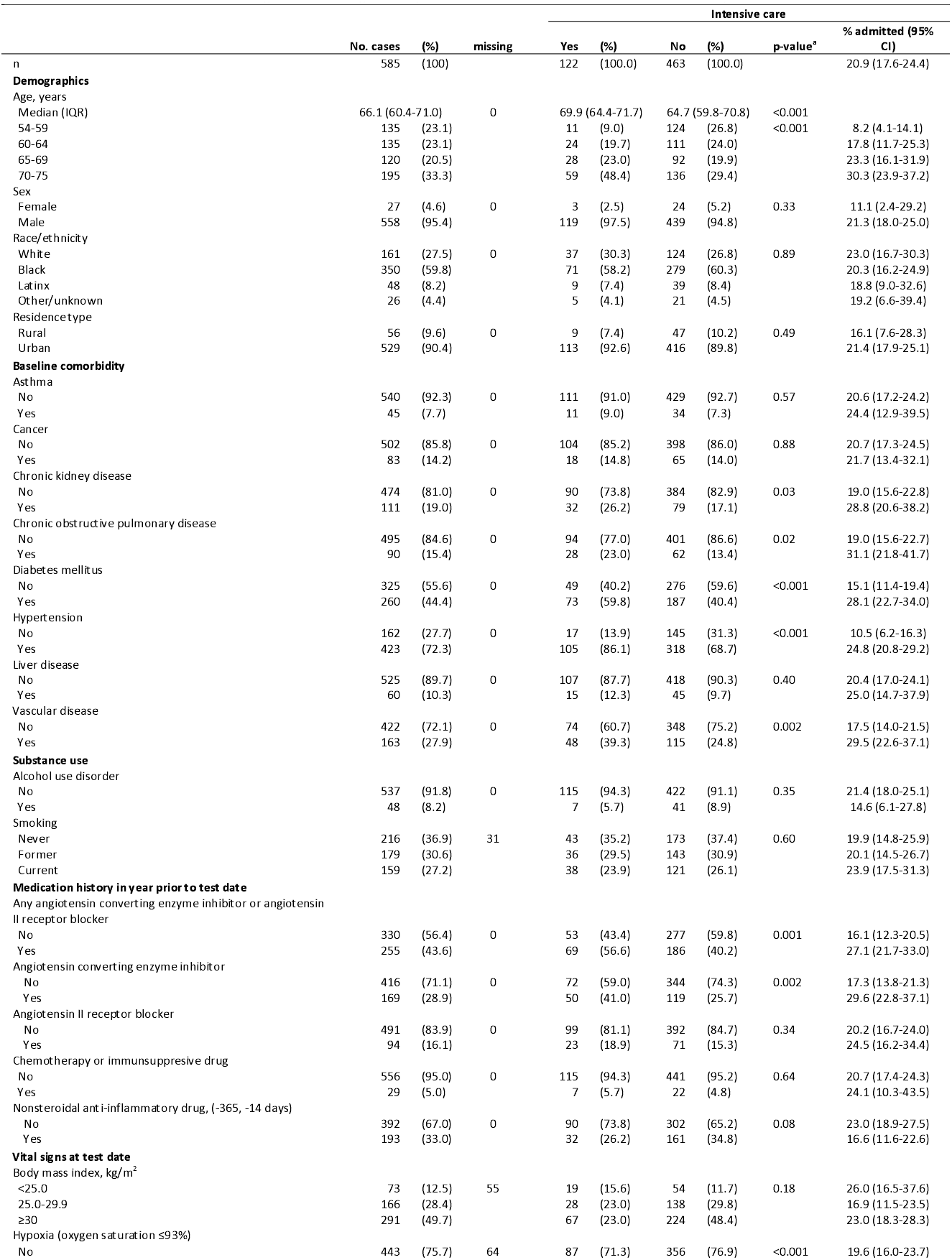
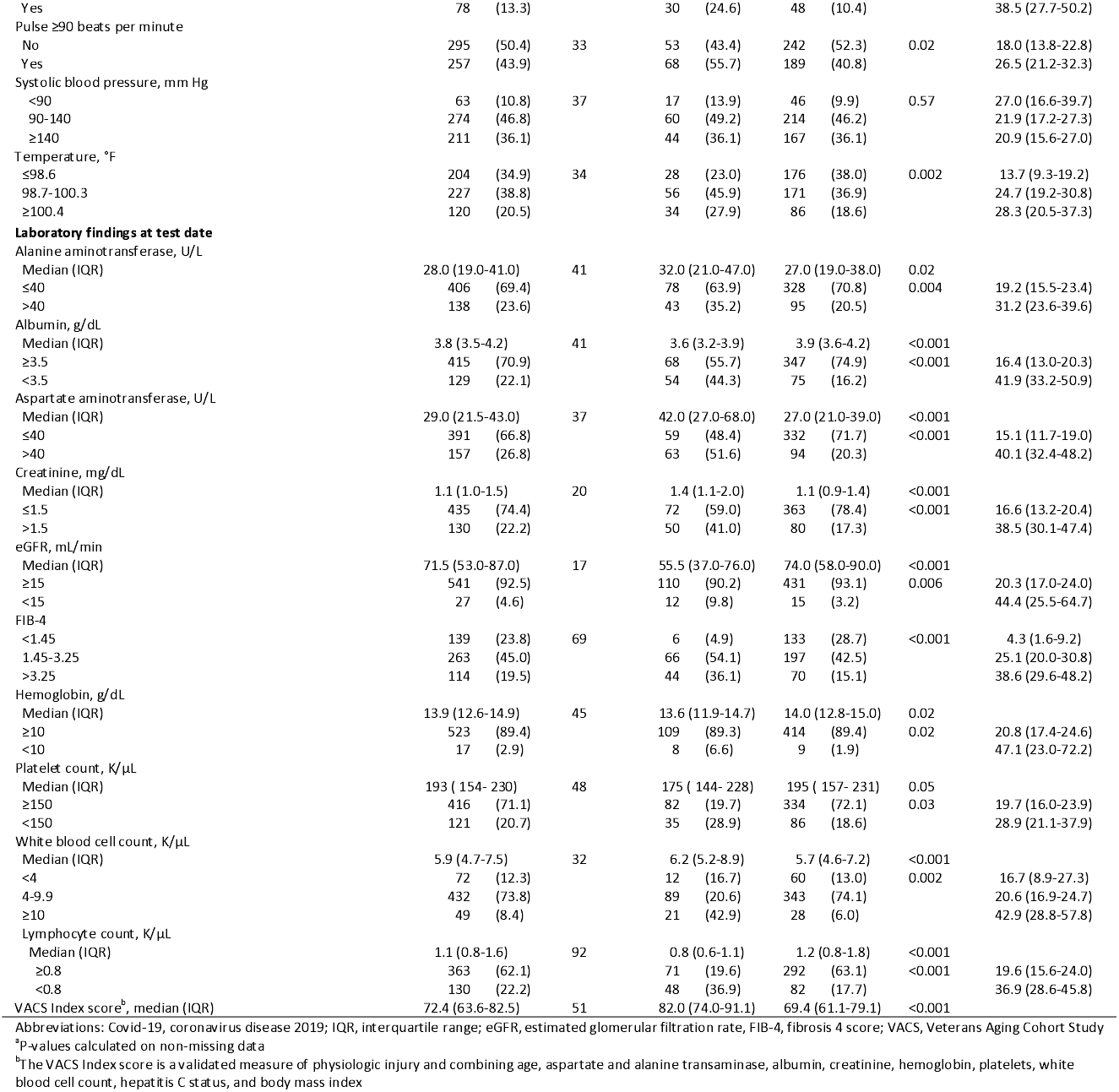
Associations with intensive care among Covid-19 cases aged 54-75 years in the Veterans Health Administration as of March 30, 2020

Parallel models, first adjusted for all significant factors identified in bivariate analyses and then substituting VACS Index for all laboratory tests, demonstrated good discrimination for hospitalization (**Table 4**, *C*-statistics: 0.859, 0.834) and intensive care (*C*-statistics: 0.876, 0.835). White blood cell count, lymphocyte count, eGFR, albumin and FIB-4 were all independently associated with hospitalization and intensive care (**Table 4**). The most pronounced association was for patients with FIB-4>3.25 – adjusted OR 8.73 (95% CI 4.11-18.56) for hospitalization and 8.40 (95% CI 2.90-24.28) for intensive care – compared to those with FIB-4<1.45. Of note, associations were stronger for FIB-4 and eGFR than for components of these measures (data not otherwise shown).

**Table 4.** Crude and adjusted associations with hospitalization and intensive care among Covid-19 cases as of March 30, 2020

|  | Hospitalization (n=297/585) |  |  | Intensive care (n=122/585) |  |  |
| --- | --- | --- | --- | --- | --- | --- |
|  | Univariable OR<br>(95% CI) | Multivariable<br>OR (95% CI) | Multivariable OR<br>(95% CI) | Univariable OR<br>(95% CI) | Multivariable<br>OR (95% CI) | Multivariable OR<br>(95% CI) |
| **Model details** |  |  |  |  |  |  |
| C-statistic | - | 0.859 | 0.834 | - | 0.876 | 0.835 |
| **Demographics** |  |  |  |  |  |  |
| Age, per 5-year increase | 1.26 (1.10-1.44) | 0.87 (0.71-1.05) | 0.64 (0.51-0.80) | 1.55 (1.30-1.86) | 1.31 (1.03-1.66) | 0.98 (0.76-1.26) |
| Race/ethnicity, black vs. other | 1.30 (0.94-1.82) | 0.96 (0.61-1.53) | 1.14 (0.74-1.74) | 0.92 (0.61-1.38) | 0.94 (0.55-1.63) | 0.98 (0.60-1.61) |
| **Baseline comorbidity** |  |  |  |  |  |  |
| Chronic kidney disease, yes vs. no | 2.36 (1.53-3.66) | 0.80 (0.41-1.56) | 0.84 (0.47-1.49) | 1.73 (1.08-2.77) | 0.50 (0.24-1.06) | 0.65 (0.35-1.21) |
| Chronic obstructive pulmonary disease, yes vs. no | 2.31 (1.43-3.72) | 1.77 (0.92-3.41) | 1.15 (0.62-2.12) | 1.93 (1.17-3.18) | 1.81 (0.92-3.58) | 1.47 (0.79-2.73) |
| Diabetes mellitus, yes vs. no | 1.96 (1.41-2.73) | 1.59 (1.00-2.53) | 1.36 (0.89-2.09) | 2.20 (1.46-3.30) | 1.69 (0.96-2.98) | 1.41 (0.85-2.35) |
| Hypertension, yes vs. no | 2.25 (1.55-3.26) | 1.39 (0.78-2.46) | 1.50 (0.88-2.54) | 2.81 (1.63-4.88) | 1.51 (0.70-3.28) | 1.89 (0.93-3.87) |
| Vascular disease, yes vs. no | 2.92 (1.99-4.29) | 1.58 (0.93-2.69) | 1.58 (0.96-2.60) | 1.96 (1.29-2.99) | 0.73 (0.40-1.33) | 0.77 (0.44-1.34) |
| **Medication history in year prior to test date** |  |  |  |  |  |  |
| Angiotensin converting enzyme inhibitor or<br>angiotensin II receptor blocker, yes vs. no | 1.63 (1.17-2.27) | 1.15 (0.71-1.87) | 1.24 (0.79-1.95) | 1.94 (1.30-2.90) | 1.66 (0.94-2.93) | 1.69 (1.01-2.84) |
| Nonsteroidal anti-inflammatory drug, yes vs. no | 0.78 (0.55-1.10) | 1.18 (0.74-1.89) | 1.09 (0.71-1.67) | 0.67 (0.43-1.04) | 1.16 (0.65-2.06) | 0.98 (0.57-1.67) |
| **Vital signs** |  |  |  |  |  |  |
| Systolic blood pressure, per 5 mm Hg decrease | 1.10 (1.03-1.17) | 1.08 (1.00-1.18) | 1.09 (1.00-1.17) | 1.05 (0.98-1.13) | 1.00 (0.92-1.10) | 1.00 (0.91-1.09) |
| Oxygen saturation, per 1% decrease | 1.14 (1.06-1.23) | 1.04 (0.95-1.14) | 1.12 (1.02-1.22) | 1.16 (1.09-1.24) | 1.05 (0.97-1.14) | 1.13 (1.05-1.21) |
| Pulse, per 5-beats per minute increase | 0.97 (0.95-0.99) | 0.97 (0.95-0.99) | 0.97 (0.95-0.99) | 0.99 (0.97-1.00) | 1.00 (0.97-1.02) | 0.99 (0.97-1.01) |
| Temperature, per 1°F increase | 1.20 (1.07-1.36) | 1.15 (0.98-1.34) | 1.24 (1.08-1.44) | 1.28 (1.12-1.47) | 1.32 (1.10-1.58) | 1.37 (1.16-1.61) |
| **Laboratory findings** |  |  |  |  |  |  |
| Albumin, per 1 g/dL decrease | 10.43 (5.70-19.08) | 3.75 (1.91-7.35) | - | 6.17 (3.62-10.52) | 3.34 (1.70-6.54) | - |
| eGFR, per 10 mL/min decrease | 1.29 (1.19-1.40) | 1.18 (1.05-1.32) | - | 1.31 (1.21-1.43) | 1.29 (1.15-1.46) | - |
| FIB-4 |  |  |  |  |  |  |
| <1.45 | ref | ref | - | ref | ref | - |
| 1.45-3.25 | 3.92 (2.53-6.08) | 2.96 (1.69-5.17) | - | 6.31 (2.74-14.50) | 4.59 (1.72-12.22) | - |
| >3.25 | 12.37 (6.73-22.72) | 8.73 (4.11-18.56) | - | 12.43 (5.24-29.50) | 8.40 (2.90-24.28) | - |
| Hemoglobin, per 1 g/dL decrease | 1.55 (1.32-1.83) | 1.16 (0.93-1.43) | - | 1.33 (1.14, 1.54) | 1.00 (0.81-1.23) | - |
| White blood cell count, per 1 K/μL increase | 1.05 (0.96-1.15) | 1.24 (1.08-1.43) | - | 1.22 (1.10-1.36) | 1.49 (1.27-1.73) | - |
| Lymphocyte count, per 1 K/μL decrease | 2.68 (2.00-3.60) | 2.38 (1.68-3.39) | - | 2.76 (1.87-4.09) | 2.65 (1.68-4.18) | - |
| VACS Index score<sup>a</sup>, per 5-point increase | 1.48 (1.36-1.61) | - | 1.62 (1.43-1.84) | 1.45 (1.33-1.58) | - | 1.47 (1.31-1.65) |

Abbreviations: Covid-19, coronavirus disease 2019; OR, odds ratio; CI, confidence interval; eGFR, estimated glomerular filtration rate; FIB-4, fibrosis 4 score; VACS, Veterans
Aging Cohort Study
<sup>a</sup>The VACS Index score is a validated measure of physiologic injury combining age, aspartate and alanine transaminase, albumin, creatinine, hemoglobin, platelets, white blood cell count, hepatitis C status, and body mass index

While COPD, diabetes, hypertension, kidney disease, vascular disease and exposure to ACE/ARB exposure were associated with hospitalization and intensive care in unadjusted analyses, they were not significantly associated after adjusting for laboratory abnormalities and vital signs (**Table 4**). Results were similar in complete case analysis (**eTable 3**).

## Discussion

Our analysis represents over 2 million veterans, aged 54-75 years, receiving care in the largest integrated healthcare system in the United States. The study was conducted within an established cohort and based on well annotated national electronic health record data, enabling a rapid and reliable analysis of Covid-19 testing and initial outcomes. As a result, we were able to validate and extend previous findings, to include a careful consideration of who is Covid-19+ and, given a positive test, what factors were independently associated with hospitalization and intensive care. We found that black Veterans were twice as likely to be tested and 2.5 times as likely to test positive than non-black Veterans, even after adjusting for urban residence and conditioning on geographic location. While we saw modest evidence of an association between exposure to NSAIDs and risk of Covid-19+, vital signs and laboratory measures better characterized risk of hospitalization and intensive care than did comorbid diagnoses or prior medication exposures.

In unadjusted analyses, black Veterans were over four times as likely to test positive compared to non-black Veterans; adjusting for urban versus rural residence did not change this association. While black Veterans were much more likely to be tested at high prevalence facilities, conditioning our analysis by site did not eliminate the association; black race retained over a two-fold increased risk for testing Covid-19+. Of note, black Veterans were also more likely to be tested, which could dilute the proportion positive. Further, black Veterans did not experience higher rates of hospitalization or intensive care. Based on prior experience with 1918 Spanish Flu and 2009 H1N1 epidemic, public health experts have warned that minority populations may be at higher risk of infection due to reduced capacity to implement physical distancing.^21,22^ Our findings may be an underestimate of the US population as racial health disparities in VA tend to be smaller than in the private sector.^23^

Women were more likely to be tested for Covid-19 than men, and men were twice as likely to test positive. This association strengthened after adjustment and in conditional analyses (**Table 2**) but should be considered preliminary given limited numbers of women in this analysis.

As reported previously^24^ elevated temperature was independently associated with testing positive, hospital admission and intensive care, underscoring the value of including fever in the current testing algorithms. Findings from the multivariable regression suggest that other factors might also be used to indicate a test, including black race, male sex, and lack of an alternative explanation for cough symptoms. To wit, we found that current smoking, COPD, and alcohol use disorder, factors that generally increase risk of pneumonia, were associated with decreased probability of testing positive. While they were not associated with hospitalization or intensive care, it is too early to tell if these factors are associated with subsequent outcomes such as respiratory failure or mortality.

Presence of particular comorbid diagnoses may be less prognostic than overall acute on chronic injury reflected in laboratory abnormalities largely encompassed in the VACS Index. In unadjusted analyses, several comorbid conditions were associated with hospitalization and intensive care but were not independently associated after adjusting for vital signs and laboratory data. Further, while elevated white blood cell counts and decreased lymphocyte counts were associated with hospitalization and intensive care, the pronounced independent association with FIB-4 (a composite of platelets and transaminases) and albumin suggest that virally induced hepatic inflammation may be a harbinger of the cytokine storm.^25-27^

VACS Index, which includes FIB-4, albumin, and white blood cell count, is predictive of mortality in many clinical settings.^18^ A five-point difference in score corresponds to a 30% difference in mortality. The 12.5-point difference in medians between the Veterans who were and were not admitted, and the 12.6-point difference between those who received and did not receive intensive care underscores the wide range of prognoses seen with Covid-19. Future work will need to determine whether VACS Index might be used in medical triage of Covid-19+ patients.

Our analysis is one of the first to address concerns regarding exposure to NSAIDS and ACE/ARBs and Covid-19.^9,10^ We found NSAID exposure was modestly associated with Covid-19+ in unadjusted and adjusted analyses, but not with hospitalization or intensive care. Among those testing positive, ACE/ARB exposure was associated with hospitalization and intensive care in unadjusted analyses, but associations lost statistical significance with hospitalization and diminished with intensive care after adjusting for clinical measures, including hypertension and blood pressure. However, confidence intervals were wide, include clinically important differences, and conclusions may change as the epidemic evolves. We will continue to update these analyses as more data become available.

While this analysis adds information to the evolving pandemic, its limitations must be kept in mind. First, a small proportion of Veterans have been tested and rates of testing vary widely by site. Second, women represented a small number of Veterans in the sample (184 tested, 13 positive). Third, our analysis of outcomes is preliminary as many Covid-19+ patients are still in care. Fourth, while a strength of this analysis is our ability to determine active VA medications, we could only detect NSAID exposure based upon VA pharmacy fill/refill data, individuals are also likely to purchase NSAIDS over the counter. As real-world data become available, more sophisticated and focused pharmacoepidemiological analyses will be required to address concerns regarding potential risk of medications associated with Covid-19.

## Conclusion

Black race was strongly associated with Covid-19+, but not with hospitalization or intensive care. Unadjusted associations between medication exposure, comorbid disease, and hospitalization and intensive care are diminished after adjustment. Risk of hospitalization and intensive care associated with Covid-19 may be better characterized by vital signs and measures of physiologic injury than by comorbid conditions or medication history.

## Data Availability

The analytic data sets used are not permitted to leave Veterans Affairs Healthcare System firewall. This limitation is consistent with the authors’ past work and with other studies based on Veterans Affairs Healthcare System data.

## Acknowledgements

The views and opinions expressed in this manuscript are those of the authors and do not necessarily represent those of the Department of Veterans Affairs or the United States Government. The authors wish to recognize Dr. Kendall Bryant as the NIAAA Scientific Collaborator for the Veterans Birth Cohort. The authors thank Dr. Jennifer Thompson for her feedback regarding helpful data presentation and important characteristics to include to enable statistical modeling of the Covid-19 pandemic.

